# Engagement with daily testing instead of self-isolating in contacts of confirmed cases of SARS-CoV-2: A qualitative analysis

**DOI:** 10.1101/2021.05.25.21257644

**Authors:** Sarah Denford, Alex F. Martin, Nicola Love, Derren Ready, Isabel Oliver, Richard Amlôt, Lucy Yardley, G. James Rubin

## Abstract

**Introduction:** In December 2020 and January 2021 Public Health England (PHE) with NHS Test and Trace conducted a study to explore the feasibility and acceptability of daily testing as an alternative to self-isolation following close contact with a confirmed COVID-19 case. This qualitative paper aims to identify factors influencing uptake among those offered daily testing, and the subsequent impact on behaviour.

**Methods:** We conducted in-depth interviews with 52 participants who had taken part in the feasibility study. Participants were asked about their experiences of daily testing or self-isolating, their reasons for choosing to test or isolate, and their behaviour during the study period. Data were analysed using inductive thematic analysis.

**Results:** Results are presented under two main headings: 1) factors influencing acceptance of testing and 2) impact of test results. Participants appeared highly motivated to engage in behaviours that would protect others from the virus. Factors influencing the decision to accept testing included 1) needing to avoid self-isolation 2) concerns about test sensitivity and 3) perceived benefits of detecting infection. Participants who were taking tests reported:1) positive consequences following confirmation of COVID status 2) engaging in essential activities 3) uncertainty and 4) self-isolating whilst testing.

**Conclusions:** This study has identified a range of factors that appear to influence the decision to engage in daily testing or to self-isolate following close contact with a positive case, many of which could be addressed by clear communications. Covid-19 infection rates and government restrictions influenced experiences, and so further research is needed to explore perceptions of daily testing and behaviour following close contact with a positive case among a wider range of individuals, in the context of lower rates of COVID-19, few government restrictions on general population behaviour and more widespread testing.

## Introduction

In the UK, a key strategy to limit the spread of SARS-CoV-2 is the requirement that, following contact with a confirmed case of COVID-19, people must self-isolate at home for 10 days. This can place a substantial burden on people, with negative impacts reported on mental health, financial strain and reduced access to education [1, 2]. For some people, particularly those who work in public-facing positions, this policy can also lead to multiple self-isolation periods [3]. Isolation of contacts can also place a burden on organisations if multiple members of the workforce are required to self-isolate at the same time, presenting significant challenges to front-line sectors, such as emergency and health care services, and education settings [1].

From the end of 2020, lateral flow device (LFD) antigen tests have become increasingly used in a variety of settings, including schools and businesses, to provide a rapid assessment of the presence of infection [4-6]. Between December 2020 and January 2021, a study was carried out by Public Health England (PHE) with NHS Test and Trace to explore the feasibility and acceptability of daily testing as an alternative to self-isolation following close contact with a confirmed COVID-19 case [7]. Where people accepted home-testing, they were asked to perform an LFD test at home once a day for up to six days. If each test was negative they could continue to leave their home, within the limits of local guidelines [8]. Details of the feasibility study have been reported elsewhere [7, 9]. In brief, it found that daily testing is potentially acceptable to some but not all, may facilitate sharing of close contacts, and promote adherence to isolation rules.

Several uncertainties still exist about implementation of this system, however, which need to be addressed before it can be rolled out more widely. Some of these relate to how willing people are to carry out home testing as an alternative to self-isolating, how people carry out and interpret their tests, and how they behave following a positive and negative test result. In this study, we carried out in depth interviews with individuals who were offered home-testing as an alternative to self-isolation following close contact with a confirmed case, as part of the previous PHE / NHS Test and Trace study. We over-sampled individuals from ethnic minority backgrounds and those who declined testing, as these groups were under-represented in the original study analysis. The aim of our study was to gain a better understanding of factors influencing acceptance of home testing, and for those who accepted, to explore the impact of test results.

## Method

We interviewed participants who took part in a study carried out by PHE with NHS Test and Trace exploring the feasibility and acceptability of daily testing as an alternative to 10 days self-isolation following close contact with a positive COVID-19 case. Full details of the methods of that study are reported elsewhere [7, 9]. In brief, between 11 and 23 December 2020 and 4 to 12 January 2021, asymptomatic contacts of confirmed Covid-19 cases were given the option to carry out LFD antigen tests at home, as an alternative to self-isolation. Those who consented were asked to complete up to six tests and report results daily using an online system. If a test result was negative, the individual could continue with daily activities for the next 24 hours, including leaving the home, within the limits of governmental policy restrictions in place in their local area [8]. The study received ethical approval from Kings College London’s Psychiatry, Nursing and Midwifery Research Ethics Subcommittee: Reference HR-20/21-21592 and Public Health England’s Research Ethics and Governance Group: Reference NR0235.

### Recruitment and data collection

We used purposive opportunity sampling to recruit participants who were offered home testing and either accepted or declined. We targeted participants from ethnic minority groups, and those who declined home testing, as these were under-represented in the original study analysis [7, 9] and we considered their views particularly important for informing efforts to maximise acceptability, feasibility and inclusivity of daily testing. All participants who we approached for interview had provided additional consent to be contacted by our team. All were 18 years or older. Additional verbal consent for our study was obtained prior to the interview.

We asked participants about their experiences of home testing or self-isolating, with a focus on their motivation for their chosen option and their experiences and behaviour during home testing or self-isolation. Full interview schedules are reported in Supplement 1. Interviews were audio-recorded and transcribed verbatim. Following the six stages of thematic analysis [10], the two lead authors independently assigned initial codes to the interview transcripts using NVivov12 software. The research team met regularly to discuss the codes and agree a preliminary set of themes. As analysis progressed, this list was refined, and similar themes were combined. Charts were developed for each theme, and verbatim quotes for each theme were collated. Charts were then used to identify narratives within and between cases.

## Results

Fifty-two participants took part in the interviews, including 17 who declined the offer of home testing and opted to self-isolate instead. Participants were aged between 18 and 73 years (mean 43 years). Thirty-three participants were female, and 18 were from ethnic minority groups (Table 1).

**Table 1:**
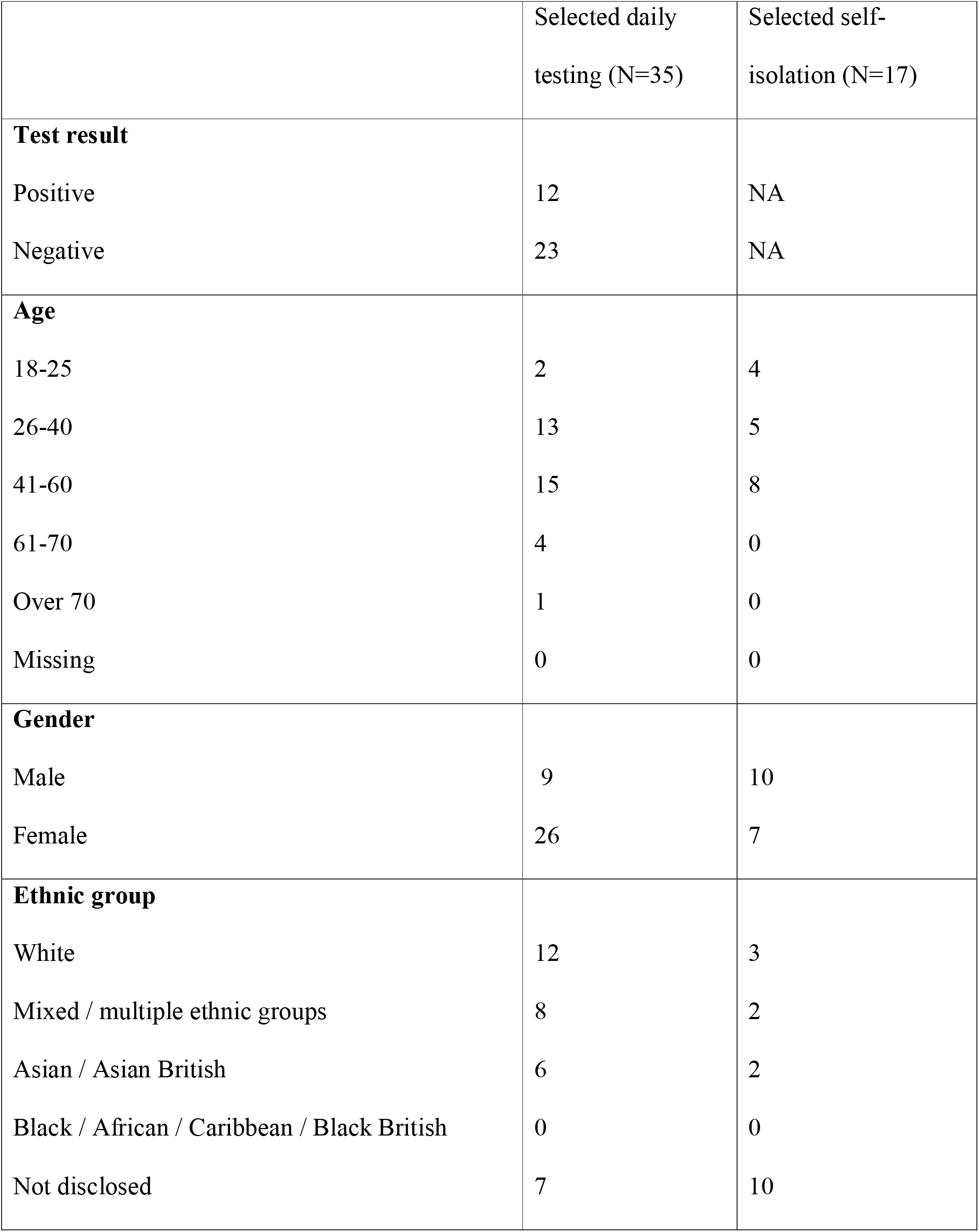
Participant characteristics

### 1. Factors influencing acceptance of testing

#### Protecting those around you

Regardless of whether participants chose to take daily tests or self-isolate, the need to protect themselves and those around them was a priority:

*“You don’t want to put anybody else at risk, so you don’t do anything that you could pass onto anybody else” (female, mixed ethnicity, consented to testing)*

However, whilst participants were highly motivated to do all they could to protect themselves and others around them from COVID-19, there was a conflicting need to minimise the impact of self-isolation on their lives. This resulted in participants weighing up the level of risk associated with various behaviours against their need to perform various activities:

*“You’re really fed up of being inside, but at the same time you think if there’s a chance that I could be going out too early and spreading it people, of course you don’t want to do it. It was a difficult call”(female, white, self-isolated)*

#### Need to avoid self-isolation

There appeared to be considerable variation in the extent to which participants needed, and wanted, to avoid self-isolation and leave their homes. Participants explained how they were willing to take daily tests to enable them to perform essential activities, such as shopping, collecting medication, and for daily exercise. Due to the festive period, the ability to run errands appeared to be particularly important, especially for those who were unable to secure supermarket deliveries:

*“[If] I were negative then I could go about, at least nip out and go to the shops just for some vital items”(male, white, consented to testing)*

*“We were just coming up to Christmas. Of course… and I hadn’t got any Christmas shopping. I thought, Oh My God. Knowing where we live, you just cannot get online shopping”(female, mixed ethnicity, consented to testing)*

Exercise was another key driver for people who had limited or shared space:

*“I’m in a Uni house, I’ve got a very small room, so for me being able to go out and go for a walk was a big thing”(female, white, consented to testing)*

For participants who needed to complete essential activities, such as collecting medication, the ability to leave the house caused considerable relief:

*“The pharmacy couldn’t deliver to us, we couldn’t do anything, so I was like how am I going to get this? Am I going to wait round until ten days just to get this prescription for my [relative]? I think in that way it was just a huge stress release to know that I can get out and get it” (female, ethnicity not provided, consented to testing)*

Whilst motivation to avoid self isolation generally led people to take part in daily testing, for some, the strong need to avoid isolation reduced their willingness to take tests. This was due to concerns about potentially extending the isolation period:

*“I had a conversation with my work shortly after and said, ‘Look, I’ve been offered this seven-day trial where if you’re negative test you can go out for 24 hours, but if in that seven days I get a positive test result then I have to do the ten days from then*.*’ I think they then said, ‘No, I think it’d be better if you stick to the ten days,’ so I just stuck to the ten days…. worst case scenario is if you’ve been isolating for nine days and then you get a positive test result and then have to do a 17-day isolation” (male, ethnicity not specified, self-isolated)*. In contrast, a number of participants explained how a lack of need to go out – even for essential tasks – resulted in them opting to self-isolate:

“*To be honest with you, working in the home, we’ve pretty much done isolation since this all began nearly a year ago” (female, ethnicity not specified, self-isolated)*.

#### Concern about test sensitivity

For some participants, doubts about the accuracy of LFD tests led them to choose to isolate rather than home test.

“*That test, I wasn’t sure of, because I understand that sometimes, even if you are positive, it can come back as negative, because I work in the NHS and I use it as well. Because I work with vulnerable people, I didn’t want to put them in risk. That is the reason why I thought it would be better to self-isolate rather than going to work” (male, Asian, self-isolated) “I think I would if I didn’t have my job. I think I definitely would [consent to testing] if I didn’t have my job, because then I won’t have anything on my conscience”(female, mixed ethnicity, self-isolated)*.

Some participants were concerned that they would potentially put others at risk if they had not taken the test accurately.

*“I didn’t feel confident that I would have been doing it properly, if that makes sense… Because I think, you know when you have to swab yourself, you tend to chicken out sometimes, if that makes sense? Not that you’d do it on purpose… But you’ve got to think you could be carrying the virus, whether you’ve got symptoms or not now, haven’t you?”(female, mixed ethnicity, self-isolated)*

Concerns about the safety of daily testing appeared to be related to participants’ concerns about the potential for transmitting the virus. One participant described a concern that he would be able to transmit the virus – even if he did not have it himself:

*“I did not want to be a carrier in any form. I mean, not necessarily within my body, but what if I carried the virus on my breath or some part of me carried it, then I went out it could spread to other people, so that’s why”(male, Asian, consented to testing)*

Participants were also worried that they would be able to contract and transmit the virus within the 24 hour window between tests:

*“If I tested negative and I went out and then the next I tested positive, I’d be thinking to myself, was that test yesterday positive and I went out and infected people?” (female, white, consented to testing)*

This was particularly true among people who were living with a positive index case, as one participant explained that he would not be willing to take daily tests due to his belief that he would, at some point, catch the virus from his partner:

*“I decided to carry out the quarantine rather than daily testing because for me, if it’s an airborne virus then it doesn’t make sense if I’m still sleeping next to the person who’s been told they’re positive for me not to have it*.*”(male, ethnicity not specified, self-isolated)*

#### Perceived benefits of detecting infection

In contrast to those voicing concerns about the transmission risks of daily testing, some participants described being motivated to take part in daily testing to make sure that they had not caught, and were not spreading the virus:

*“The reason I decided [to take tests is] because my family caught Covid… They contracted Covid and when I was offered these tests, I wanted to make sure that I take them on a daily basis and make sure I don’t get Covid and basically preventing the spread of Covid” (male, Asian, consented to testing)*

Participants described how testing could help protect vulnerable people in their household: *“I thought it was a great addition that I could have that resource available so that I knew that the vulnerable people in my house wouldn’t be put in any harm” (female, ethnicity not specified, consented to testing)*

One participant described how testing could facilitate self-isolation and so prevent spread of the virus:

*“Also, it’s good to know whether you’ve got it or not, so that you then know that you have to isolate so that you’re not transferring it around” (female, mixed ethnicity, consented to testing)*.

### 2. Impact of test results

#### Positive consequences of confirmation of COVID status

Participants who went on to develop symptoms during the study, described how tests provided rapid confirmation of their COVID status:

*“When I first felt ill, I didn’t immediately think it was Covid; I was convincing myself it wasn’t. I was like, ‘Well, I haven’t got a temperature, and I haven’t got a cough…’ Although I did get a cough, eventually. I was like, ‘No, it’s not going to be Covid. It’s not, it’s not; it’s just the flu*.*’ No, actually, it was only from having a positive test at home that I knew” (female, mixed ethnicity, consented to testing)*

Tests were often used in combination with (a lack of) symptoms as a way of providing additional reassurance that they did not have the virus:

*“You blow your nose, you sneeze, you think, aye-aye, and then you start to get paranoid. I eat like a horse anyway and I can smell a fart from about 300 yards, so I know there’s no trouble with me, pardon the expression” (male, white, consented to testing)*

Participants described how it helped their mental wellbeing to know whether or not they had the virus:

*“It’s just that peace of mind because I was genuinely having sleepless nights, worrying, but I wouldn’t tell my [relative] that. The fact that it was in the same house and the [relative] told me, that they said it could be fatal to me, was a worry. So it was lovely to have that reassurance that every morning” (male, white, consented to testing)*

Participants were not always concerned about having a positive test result, but were eager to know either way:

*“When I did the tests and I got the results at the end of it within 30 minutes, it gave a peace of mind that obviously, that I was not positive on the days, or I was… Either I was positive or negative, within the 30 minutes I would know that, so it was ease of finding out where I am with Covid and basically, getting the results at such a pace, it just gave us peace of mind more than anything else” (male, white, consented to testing)*

#### Enabling essential activities

Several participants described how negative test results enabled them to undertake essential activities, such as collecting medications:

*“When I was negative, and I thought I need to collect my [relative’s]medications and just get on with my life. When I got it I was like, let’s just do this”(female, ethnicity not provided, consented to testing)*

Others appreciated being able to spend time with loved ones over the festive period:

*“The following days, obviously the next day would have been Christmas Day, so it was nice to know that I could actually spend that day with my family… Me and my partner don’t live together, so it was nice to know that I could still meet up with him outside over Christmas because we’ve spent quite a lot of this year apart, so I think we both would have found that quite difficult” (female, ethnicity not specified, consented to testing)*

Participants appreciated being able to leave their homes for a short while, but described making efforts to ensure that they did so safely:

*“I was still fairly restrictive. To be honest, I was only popping to the supermarket, not every day. Probably out of that ten-day period I visited the shops three times, and that was only one supermarket for a short period of time. Then just daily dog walks, which again, were, living fairly rurally so we weren’t seeing any people anyway, so it was just to have the freedom to be able to do those essential things really” (female, white, consented to testing)*

*“I think I went into the supermarket once to get some bits, but other than that I didn’t do anything. Like I said, I think you can trust the result to a point, but I was still well aware that there was Covid in our house, so just tried to be responsible” (female, ethnicity unspecified, consented to testing)*

A number of participants described being particularly cautious in their behaviour during the testing period, but felt on reflection that they could have done a bit more:

*“Looking back at it now I felt like I could have done a little bit more, but I was just being precautionary. I only was still going out where I needed to go because for me, it was like even though it’s a negative I felt like I still could get it the next day. Even though the test was negative that day, that’s not to say that the next day it would be negative. I think that I just felt that I needed to take those extra steps just for my own peace of mind and to make sure that they would be safe”(female, ethnicity not specified, consented to testing)*

However, for many, the ability to leave their home was considered important for their mental health:

*For my mental health, it really did benefit me being able to know that I could go about my daily routine rather than… To stay isolated on your own for ten days, it’s not good for anyone’s mindset. Yes, I found it really good to have that option available to us” (female, ethnicity unspecified, consented to testing)*

#### Uncertainty

A small number of participants who consented to home-testing were confused by the rules, and questioned whether or not they were actually allowed to leave the home.

*“I did end up phoning NHS Direct also, because I felt, like we said before, a little bit uneasy that the rules… I just wanted to double check that I was okay. I did phone NHS… They said if I had the information letter about participating in a study, and I had that with me, that I was okay to be out, and because I’d uploaded my tests electronically, so I had the test results and everything there to hand… but even my [relative] said to me, ‘Do you think you should be going out?” (female, white, consented to testing)*

The main source of confusion appeared to be whether participants were able to leave the house when living with a positive case. This may have been the result of a lack of clarity in the messages about this situation. For example, one participant described how she had been told (correctly) at the point of recruitment that she was able to take part even though many of her household had tested positive:

*“When the lady phoned me, she said, ‘If you test negative you can go out*.*’ I said, ‘What? Even though the rest of the family are positive?’ She went, ‘Well, if you’re negative you can go out*.*’ I thought, yes!” (female, white, consented to testing)*

Formal study documentation stated that to continue to work and visit shops “we would encourage you to separate yourself from the person (with COVID-19) in your household as much as possible during the study period”, which people interpreted in different ways: *When I received the paperwork it said that it didn’t apply if the person lived within your household. It was only if you’d been in contact with that person and had no further contact with them, so I was a bit confused as to whether or not it did actually apply to me or not”(female, ethnicity not specified, consented to testing)*

One participant who was living with a positive case was confused about whether they were able to go to work:

*“The plan was to get back to work sooner, but I was a bit confused over that because the person that I’d been in contact with lived with me… so I didn’t end up going back to work, but that was my initial reason for wanting to do it”(male, white, consented to testing)*

Those living with a positive case sometimes combined isolating with testing as a result of being confused by the rules: :

*“Well, to be honest, I did quarantine anyway because I was actually living with my [relative] at the time, so I think it was different, that I still had to quarantine even though I was doing the test kit” (female, white, consented to testing)*

However, a level of uncertainty was evident – even within families:

*“We had this big argument with, my [relative] and myself, because he said to me, ‘You can’t go out even if it says negative…*.*’*. *I said, ‘But it does say in here that if I was living on my own and I was negative I could go out*.*’ He went, ‘No, you’re reading it all wrong*.*’ So we had this discussion a couple of times, and in the end I thought, I give up. I wasn’t going out anyway” (female, mixed ethnicity, consented to testing)”*

#### Self-isolating whilst testing

Despite consenting to take daily tests, a considerable number of participants were still reluctant to leave their homes. This group described restricting their behaviour, over and above what was recommended, by still not going out despite a negative test result:

*“Even if the tests that were given to me, the rapid tests allowed me to step out for 24 hours, I decided not to basically do it. Yes, again, I just followed the basic national guidelines, etc*., *etc*., *looking at all of that, but just decided to self-isolate”(male, Asian, consented to testing)*. This group used test results to allow them to spend time with others within their household: *“They did obviously, indicate that as long as the tests were negative, I could go around my daily routine, so once that test was negative, I then was able to leave my bedroom and just… I still did kind of socially distance from my family within the house… but I was able to obviously, spend some time with them and not feel so isolated, so that was good” (female, ethnicity unspecified, consented to testing)*

However, despite initial motivations for taking part in daily testing, participants reported feeling uncomfortable when it came to leaving their home:

*“I did initially think I’ll just be able to go into uni and things like that, but I didn’t end up doing that because it didn’t quite feel right”(female, white, consented to testing)* Following a seven day testing period, one participant felt obliged to then isolate for the remainder of the 10 day isolation period:

*“I actually waited then, because it was seven days after, but I kind of waited until my ten days was up before I ended my isolation period, if you like” (female, white, consented to testing)*

## Discussion

### Main findings of this study

To our knowledge, this is the first study to qualitatively explore perceptions of daily testing as an alternative to self-isolation following close contact with a positive case. In choosing whether to take daily tests or self-isolate, participants described a decision-making process that ultimately aimed to maximise the safety of themselves and others around them, whilst minimizing any detrimental impacts of self-isolation. This involved individual assessments of the extent to which avoiding self-isolation was necessary and important, combined with level of confidence in the safety and accuracy of daily testing.

Participants varied in their need and desire to avoid isolation. In line with previous research, those who were unable to work remotely, unable to access support or supplies, or who lived in crowded accommodation with limited outside space appeared to have a greater need to avoid isolating than those who had a strong support network and were able to work remotely [11]. The decision to decline testing could stem from either a lack of need to avoid self-isolation, or in a small number of cases a strong need to avoid self-isolation. This latter group described concerns that a positive test result could potentially extend the standard 10 day isolation period – something that they were highly motivated to avoid. Pressure from employers appeared to exacerbate this; highlighting the need for support for isolation from employers. In our sample, some employers appeared to hold an irrational view of self-isolation, preferring that a potentially infectious member of staff attend work than allowing them to remain in self-isolation beyond the originally intended 10 day period. Any widespread roll-out of daily testing should include communications for employers highlighting the benefits of the system in reducing the likelihood of workplace outbreaks. Perceptions of the safety of testing appeared to be influenced by participants’ beliefs about the accuracy of tests and transmissibility of the virus. Due to media coverage and ongoing debates about the accuracy of the LFD [12] many participants indicated surprise and concern regarding their use. A number of participants, particularly those who were living with someone who had the virus, reported concerns that they could potentially transmit the virus even if they had not tested positive. This included concerns that they could carry the virus on their clothes, be infectious before they tested positive, or contract the virus within the 24 hour window. Greater clarity about the use of LFDs among people who live with a case will be required in any future roll-out. Study documentation stated that people may still leave the home following a negative LFD test, provided that they separate themselves from the positive case as much as possible but this was interpreted in different ways. Our findings also highlight the need for support for self-isolation among those who do not feel comfortable or able to continue with their normal daily routines following exposure to the virus. Test results provided participants with valuable confirmation of COVID status, particularly when faced with uncertain or unusual symptoms. For vulnerable individuals, or those highly concerned about catching the virus, confirmation provided much needed reassurance and reduced unnecessary anxiety. This finding tallies with a wider literature on the impact of emerging health risks, where uncertainty about exposure status is often cited as anxiogenic [13, 14].

In some cases, testing facilitated the identification of a positive case, leading to immediate isolation. Importantly, these participants appeared to have been struggling with their understanding of the symptoms of COVID-19 expecting either that loss of smell would be the key symptom or convincing themselves that their symptoms would not be COVID-19. In these cases, the provision of daily testing helped them to receive an earlier diagnosis. In accordance with existing literature, this suggests that a confirmatory test results is likely to facilitate adherence to social distancing measures [9].

Participants were informed that they could leave the house for 24 hours following a negative test result, and our previous work found that participants did engage in more activities following a negative test result than on the days that they were trying to self-isolate. However, only a small percentage reported engaging in more high contact activity than they had before the testing period [9]. Whilst no additional restrictions were mentioned in terms of what participants were and were not able to do during the 24 hour period, it was clear that participants did not accept the result of the LFD as conclusive evidence that they did not have the virus, and still made every effort to distance as much as possible. Indeed, many participants choosing to take daily tests to enable participation in essential activities reported that they had restricted their behaviour more than they had prior to testing, and others still engaged in self-isolation, despite negative test results. This was particularly important among those who were living with someone who had the virus. Whilst this is in line with previous research [15] that found participants engage in social distancing measures over and above what is recommended, it may be argued that those living with a positive case are more likely to catch the virus themselves, therefore cautious behaviour is sensible. This belief may explain why our previous work found that individuals receiving a positive test result reported less contact on the days that they had a negative test result than those that received negative test results throughout [9].

### Limitations

The main limitation associated with this work is the extent to which the views of our sample can be transferred to other settings and sectors of the community. Whilst every effort was made to recruit a diverse sample of individuals, it is inevitable that not all groups were included. In particular, those declining daily tests were underrepresented in the sample. Furthermore, many participants reported being able to work remotely, and/or were not financially disadvantaged as a result of having to isolate.

Secondly, many of the participants in the study reported excellent adherence to the guidelines at all times, whereas anonymous surveys have found breaches of adherence are common [16]. It is possible that participants did not feel able to admit to breaches – even if they were considered to have been necessary at the time. Participants who had poor adherence may have been less likely to take part in the study. However, our findings are in line with ONS data reporting adherence in 90% of people required to self-isolate following close contact with a confirmed case [17].

Finally, data collection for this study began just as the UK went into a third lockdown, and considerable social distancing restrictions were in place. This will have had a considerable impact on the extent to which many participants had a need to go to work or leave the home for other reasons. Future research should explore a diverse range of views in a context in which lockdown restrictions have been eased and the familiarity with testing has increased [18].

## Conclusion

This study has described the range of factors that appear to influence the decision to engage in daily testing or to self-isolate following close contact with a positive case. Participants were highly motivated to protect themselves and those around them, and engaged in a critical assessment of their own situation to ultimately decide how they could achieve this. Research must now explore perceptions of daily testing and behaviour following close contact with a positive case among a wider range of individuals, outside of lockdown, and in an environment in which cases of COVID-19 are lower and testing is more common.

## Data Availability

The datasets used and/or analysed during the current study are available from the corresponding author on reasonable request.

## Declarations

### Ethics approval and consent to participate

Ethical approval for this study was granted by Kings College London’s Psychiatry, Nursing and Midwifery Research Ethics Subcommittee: Reference HR-20/21-21592 and Public Health England’s Research Ethics and Governance Group: Reference NR0235.

### Consent for publication

All participants provided verbal consent for data to be included in publications

Availability of data and materials

### Authors’ contributions

Conceived the study: All authors

Study design: All authors

Analysed the data: AM

Interpreted the data: All authors

Drafted the manuscript: AM and SD

Reviewed the manuscript and approved content: All authors

Met authorship criteria: All authors

## Acknowledgements

Lucy Yardley is an NIHR Senior Investigator and her research programme is partly supported by NIHR Applied Research Collaboration (ARC)-West, NIHR Health Protection Research Unit (HPRU) in Behavioural Science and Evaluation, and the NIHR Southampton Biomedical Research Centre (BRC).

Sarah Denford is supported by the NIHR Health Protection Research Unit (HPRU) in Behavioural Science and Evaluation at the University of Bristol in partnership with Public Health England.

Alex F Martin is supported by the Economic and Social Research Council Grant Number ES/J500057/1 and the NIHR HPRU in Emergency Preparedness and Response at King’s College London in partnership with Public Health England.

James Rubin is supported by the NIHR HPRU in Emergency Preparedness and Response at King’s College London in partnership with Public Health England.

## References

1. 1. HM Government. Attendance in education and early years settings during the coronavirus (COVID-19) outbreak 2021. Attendance in education and early years settings during the coronavirus (COVID-19) outbreak – 23 March 2020 to 10 June 2021 - Official statistics announcement - GOV.UK (www.gov.uk) [Accessed May 2020]

2. Brooks S, Webster RK, Smith L, Woodland L, Wessely S, Greenberg N, et al. The psychological impact of quarantine and how to reduce it: Rapid evidence review. Lancet, 2020. 395: p. 912–920.

3. Rubin G, Smith L, Melendez-Torres GJ, Yardley L. Improving adherence to ‘test, trace and isolate. Journal of the Royal Society of Medicine 2020. 113(9): p. 335–358.

4. Torjesen, I., COVID-19: How is the UK using lateral flow tests in the pandemic? BMJ, 2021. 372: n287.

5. Department of Health and Social Care. More employers sign up to rapid testing to protect workforce. 2021. More employers sign up to rapid testing to protect workforce - GOV.UK (www.gov.uk) [Accessed May 2020]

6. Department of Health and Social Care. Pilot for family members to get regular testing for safer care home visits. 2020. Pilot for family members to get regular testing for safer care home visits - GOV.UK (www.gov.uk) [Accessed May 2020]

7. Love N, Ready D, Turner C, Yardley L, Rubin J, Hopkins S, Oliver S. The acceptability of testing contacts of confirmed COVID-19 cases using serial, self-administered lateral flow devices as an alternative to self-isolation. MedRxiv [preprint] (2021). Available at The acceptability of testing contacts of confirmed COVID-19 cases using serial, self-administered lateral flow devices as an alternative to self-isolation | medRxiv

8. HM Government. COVID-19 Response - Spring 2021. 2021: https://assets.publishing.service.gov.uk/government/uploads/system/uploads/attachment_data/file/963491/COVID-19_Response_-_Spring_2021.pdf. [Accessed May 2020]

9. Martin AF, Denford S, Love N, Ready D, Oliver I, Amlot R, Rubin J, Yardley J. Engagement with daily testing instead of self-isolating in contacts of confirmed cases of SARS-CoV-2. MedRxiv [preprint], 2021. Available Engagement with daily testing instead of self-isolating in contacts of confirmed cases of SARS-CoV-2 | medRxiv

10. Braun, V. & Clark, V. Using thematic analysis in psychology. Qualitative Research in Psychology, 2006. 3(2): p. 77–101.

11. Smith, L, Amlot R, Lambert H, Oliver I, Yardley L, Rubin GJ. Factors associated with adherence to self-isolation and lockdown measures in the UK: a cross-sectional survey. Public Health, 2020. 187: p. 41–52.

12. Deeks J, Raffle A, Gill M. Covid-19: government must urgently rethink lateral flow test roll out. BMJ, 2021.

13. Rubin, GJ, Amlot R, Carter H, Large S, Wessely S, Page L. Reassuring and managing patients with concerns about swine flu: Qualitative interviews with callers to NHS Direct. BMC Public Health, 2010. 10(451).

14. Rubin, GJ, Page L, Morgan O, Pinder RJ, Riley P, Hatch S, Maguire H, Catchpole M, Simpson J, Wessely S. Public information needs after the poisoning of Alexander Litvinenko with polonium-210 in London: cross sectional telephone survey and qualitative analysis. BMJ, 2007. 335(7360): p. 1143.

15. Denford S, Morton K, Lambert H, Zhang J, Smith LE, Rubin GJ, Cai S, Zhang T, Robin C, Lasseter G et al. Understanding patterns of adherence toi COVID-19 mitigation measures: A qualitaitve interview study. Journal of Public Health, 2021. fdab005.

16. Smith, LE, Potts HW, Amlot R, Fear NT, Michie S, Rubin GJ. Adherence to the test, trace and isolate system in the UK: results from 37 nationally representative surveys (the COVID-19 Rapid Survey of Adherence to Interventions and Responses [CORSAIR] study). BMJ, 2021. 372; n608.

17. Office for National Statistics. Coronavirus and self-isolation after being in contact with a postive case in England: 1 April to 10 April 2021. 2021. Coronavirus and self-isolation after being in contact with a positive case in England: 1 April to 10 April 2021 - Office for National Statistics (ons.gov.uk) [Accessed May 2021]

18. Gov.UK, Understanding laterla flow antigen testing for people without symptoms. 2021. https://www.gov.uk/guidance/understanding-lateral-flow-antigen-testing-for-people-without-symptoms [Accessed May 2021]

